# Impact of Language Models on Healthcare in Thailand: Benefits, Challenges, and Future Opportunities

**DOI:** 10.1101/2024.06.10.24308731

**Authors:** Chanakan Moongthin

## Abstract

This study explores the use of Artificial Intelligence (AI), specifically Large Language Models (LLMs) in the Thai healthcare sector, focusing on applications such as diagnosis, patient monitoring, and automated question-and-answer systems. While AI has the potential to improve diagnosis accuracy, reduce the time required for appointment, and enhance patient care, several challenges prevent widespread adoption of LLMs in healthcare, including significant computational resources required for deployment, data privacy and security concerns, and Thai language being a low-resource language. Through a comprehensive analysis of publicly available online data and literature, this study examines the current state of AI adoption in Thai healthcare, identifying key barriers to adoption and providing recommendations for overcoming these challenges, including targeted training and education for healthcare professionals, strategic government initiatives, and investments in infrastructure. By addressing these issues, Thailand can harness the full potential of AI technologies to enhance its healthcare system, ensuring better patient outcomes and operational efficiencies.

## 1 Introduction

The rapid advancement of Artificial Intelligence (AI) has transformed various industries, and healthcare is no exception. From Recurrent Neural Network to Transformer [1], LLMs can understand and respond to human natural languages with an accuracy higher than before. Sparking the potential for usage in applications such as chatbots, document summarization, and risk analysis. The integration of AI in healthcare has the potential to revolutionize the sector, improving diagnosis accuracy, enhancing patient care, and streamlining clinical workflows. Certain LLMs that have not been trained on medical data, such as GPT-4, have nevertheless been successfully utilized by medical professionals and researchers to analyze and interpret medical data, revealing their potential utility in both research and practical applications [2].

However, the adoption of AI in healthcare is not without its challenges, particularly in developing countries like Thailand. This study aims to explore the current state of AI adoption in the Thai healthcare sector, identifying the key barriers to adoption and providing recommendations for overcoming these challenges.

## 2 Opportunities and Challenges of using LLMs within healthcare in Thailand

Large Language Models currently have its pros and cons to consider when using these models in certain applications. The potentials of using LLMs in healthcare field are as follows [3]:

### 2.1 Opportunities

1. Vast Information and Knowledge: LLMs have been trained on large corpus of internet data, which include medical literature, research, papers and other relevant sources, allowing LLMs to respond and analyze certain context related to medical field.
2. Accessibility and Availability: Language models are different to human, they are mostly always online and respond at speed higher than any human can, and it is also much easier and more accessible to access LLMs than to arrange an appointment with the doctors where a lot of factors could make or break an appointment.
3. LLMs can keep learning new things: LLMs can be trained and updated with the latest medical data, ensuring that LLMs always have the latest information needed, allowing for flexibility and adaptability to improve the model over time.

Despite the potential to revolutionize healthcare for everyday usage, and the applications that LLMs can be useful for, several obstacles prevent the widespread adoption of LLMs within Thailand, such as:

### 2.2 Challenges

1. Ethical Concerns regarding Privacy and Security: Since LLMs are usually large (as the name suggest), and thus require a lot of resources to run, it is usually not feasible to deploy LLMs on-premise due to cost and power usage, which encourage people to deploy them on the cloud. This approach resulted in privacy and security concerns of relying on 3rd party to process users data, especially sensitive information such as, but not limited to, user’s diagnosis information, user’s past diagnosis, etc. Making the usage of 3rd party platforms for processing and storing sensitive data impractical. And there can be concerns related to potential liability and accountability of AI systems in healthcare that have to be addressed in order to avoid potential harm to patients and legal implications.
2. Accuracy Problems: LLMs are usually infamous for its propensity to hallucinate. Hallucinations, as defined by Berrios and Dening [4] quoted “Hallucinations are conceived of as indistinguishable from real perceptions except that there is no stimulus”, and in the case of LLMs, it means that the model will produce inaccurate or nonsensical information regarding the topics. Which can be caused by low amount of information related to the topic, bias within the base model itself such as memorization at the level of sentences, and statistical patterns of usage learned at the level of corpora [5].
3. Thai is a low-resource language: When using Language Models in Thailand, having the ability to understand Thai language and context is important. Unlike English, Thai has received a lot less attention from the NLP community. Which makes finding large corpus of Thai contents to pre-train LLMs on, a more complicated task. As such, there weren’t any Thai language models that fully captures Thai context and culture. But recently, there have been progress on creating Thai large language model from multilingual datasets such as MC4, which is the cleaned version of Common Crawl’s web crawl corpus, and Oscar, which is an open-source multilingual dataset. The work to create a better-understanding LLM for Thai culture is continuously being done under the name Typhoon [6] by SCB 10X, in which we hope can help mitigate these problems.

## 3 Existing Solutions

While several challenges hinder the adoption of Large Language Models (LLMs) in Thai healthcare, some concerns can be mitigated through innovative solutions. For instance, at the basic, giving patients a warning about the service utilizing LLMs to process patients data, giving patients a choice to consent, then privacy and security concerns can be alleviated by employing smaller language models with moderate parameter counts, and enabling the model to reference external information while responding to users. One viable option is to utilize a Thai language model, such as LLaMA-3-Typhoon-v1.5-8B or LLaMA-3-Typhoon-v1.5-8B-Instruct [6], and fine-tune it for the goal of enabling LLM to respond within the desired patterns, and not necessary for imparting knowledge, as any knowledge imparted into fine-tuning steps will result in knowledge degradation to the model [7]. Subsequently, quantization techniques such as GGUF from llama.cpp [8] can be applied to reduce the model size, allowing it to be run on lower-resource system while maintaining its level of accuracy. To further mitigate accuracy problems, during inference, a retrieval mechanism can be added to the LLM, allowing it to recall data from vector storage solutions such as ChromaDB [9] or Weaviate [10]. This approach, known as Retrieval-Augmented Generation (RAG), is preferred over fine-tuning knowledge into the base model, as RAG provides a ground-truth source of information to the LLMs [11].

## 4. Methodology

### 4.1 Objective and Methods

The objective of this review is to explore the current state of research on the utilization of Large Language Models in health-care within Thailand. Specifically, this review will assess the potential benefits, address the challenges, and implications of integrating LLMs into healthcare sector.

To conduct this review, a comprehensive search was conducted using all major electronic databases, including Google Scholar, ThaiJO. The keywords used were

- “Language Models” AND “medical” (AND “Thailand” outside ThaiJO)
- “ChatGPT” OR “GPT” AND “medical” (AND “Thailand” outside ThaiJO)
- “ChatGPT” OR “GPT” AND “medical” (In Thai language)

And retrieve relevant articles published within the past five years. The selected articles were then critically analyzed to extract key findings and insights. The search is conducted on June 9, 2024. The records are then selected using the Preferred Reporting Items for Systematic Reviews and Meta-Analyses (PRISMA) guidelines, to ensure the highest quality and transparency in reporting.

### 4.2 Criteria

The systematic review was conducted according to the Preferred Reporting Items for Systematic Reviews and Meta-Analysis (PRISMA) guidelines. We use Google Scholar and ThaiJO to search candidate publications. Filters are used when searching on Google Scholar, to enable searching for articles ordered by relevance and published after 2022. The search is conducted on June 9, 2024. The eligibility criteria involved any type of published scientific research or preprints (article, review, communication, etc.) addressing the use of LLMs that fell under the medical field categories within Thailand.

The inclusion criteria are as follows:

1. Papers must be published or reported between 2020 and 2024
2. The context of the paper is about the utilization of Large Language Model (LLMs) in healthcare within Thailand.
3. The paper must be written and published in Thai or English language.
4. The paper is original and published as journal article.

The exclusion criteria included: records that are not in Thai and English, records addressing the use of LLMs in subjects other than those related to medical field, records addressing the use of LLMs in medical field outside of Thailand, and articles from non-academic sources. We excluded 116 out of 132 screened articles, leaving us with 16 full-text articles to be assessed for eligibility.

After completing the eligibility assessment during the selection process (n=7), we then proceed to review the articles as follows.

**Figure 1:**
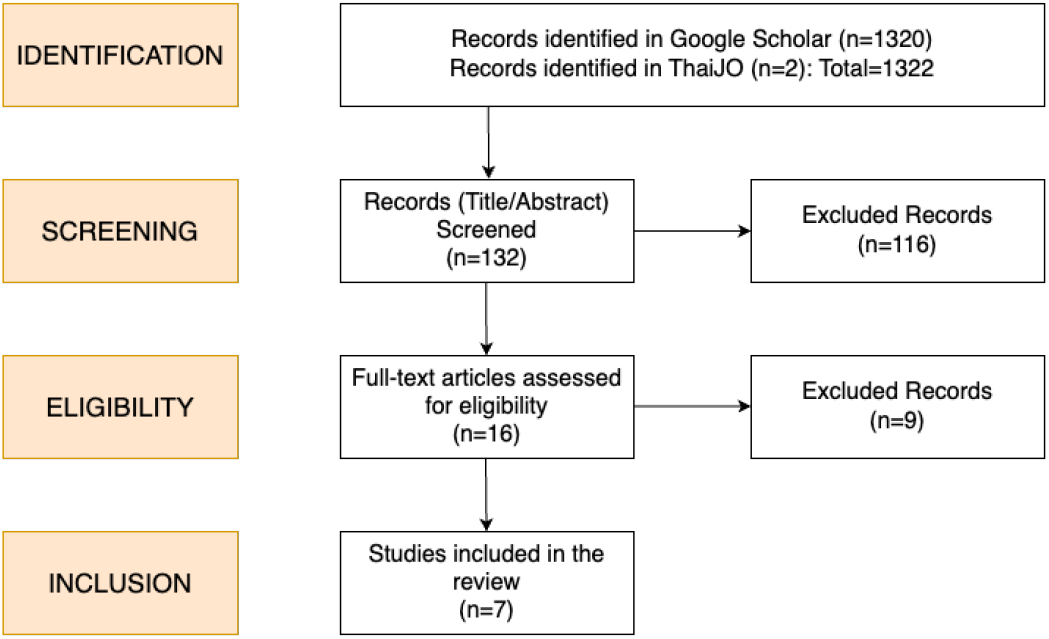
Flowchart of the record selection process based on PRISMA guidelines.

### 4.3 Results

Initially, we have a total of 2 results from ThaiJO, and 1,320 results from Google Scholar (For the first keyword).

## 5 Critical analysis of key reviews in this area

Table 1 outlines the key reviews published to date that provide examples of the LLMs applications in healthcare within Thailand.

**Table 1:**
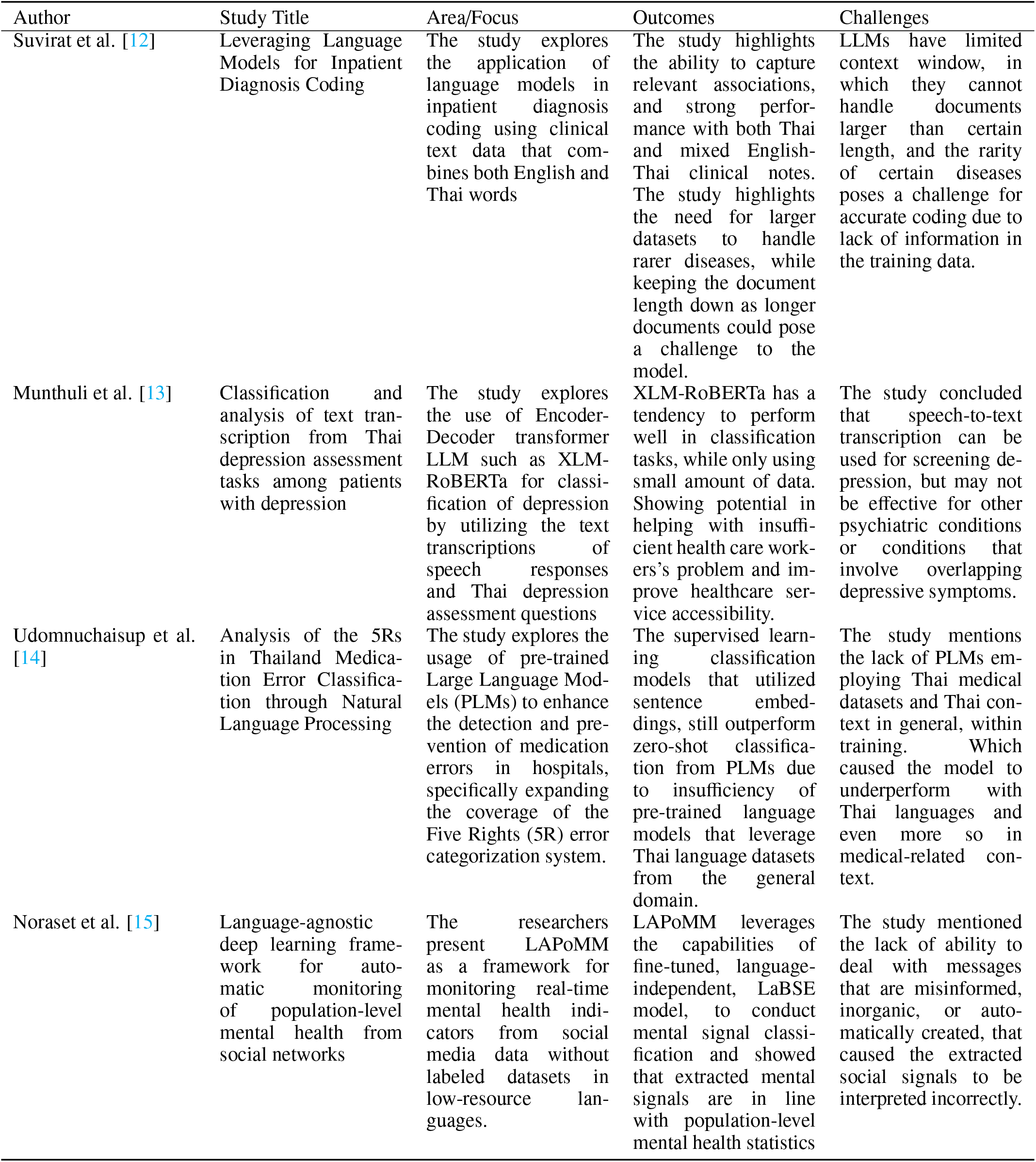
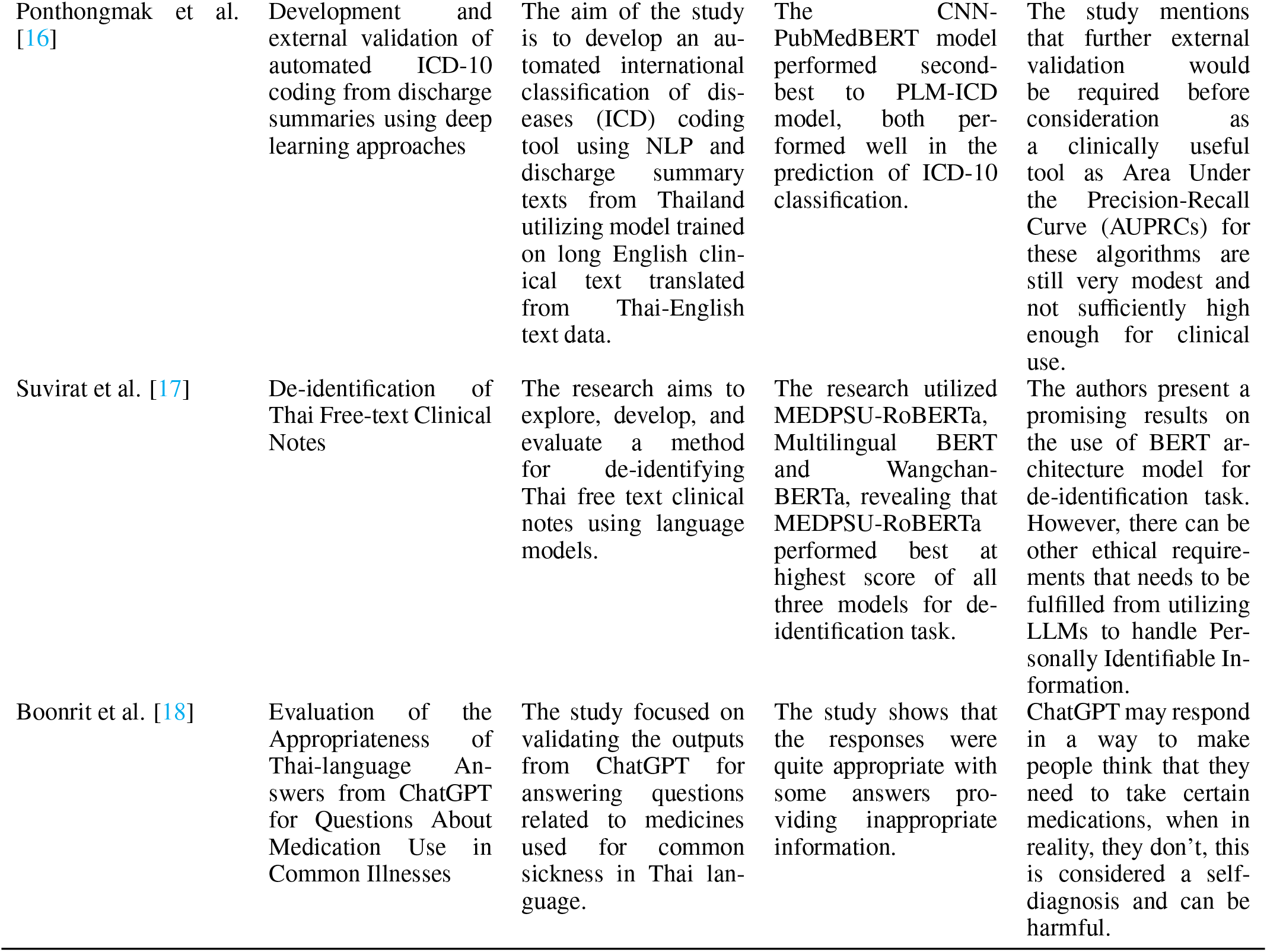
Critical Analysis of usage of LLMs application in healthcare in Thailand.

From the analysis, we found below key areas to form a generalized framework:

- Language Model Selection and Fine-tuning: The choice of language model architecture (e.g., XLM-RoBERTa) and fine-tuning on specific tasks (e.g., depression classification) can significantly impact performance, as certain pre-trained language models may not perform well on specific tasks or languages (e.g., Thai) without sufficient training data, and certain models will have different context window, so for some tasks that require processing large amount of data within one context will require language models with larger context window. Certain LLMs will be multimodal, which means the model will accept more than just text (e.g., Visual and/or Auditory) which can be helpful in certain tasks.
- Determine the usage of LLMs in certain tasks: Certain tasks such as depression screening can be done with LLMs, but certain other tasks that requires classification with overlapping symptoms may not be as accurate or useful, it is important to keep every possible parameters that can caused a specific conditions into mind.
- Data Quality and Availability: The availability and quality of training data, especially for low-resource language (e.g., Thai), can significantly impact model performance, as the lack of large, diverse datasets for specific tasks can hinder model development.
- Addressing the Rarity of Diseases and Insufficient Data: As language models work based on the data being provided, having insufficient data can hinder model performance, especially within healthcare and medical field where there may not be enough information for rarer diseases.
- Handling Misinformation and Noise in Data: Models should be designed to handle misinformation, inorganic, or automatically generated data to ensure accurate interpretations.
- Evaluate model performance on multilingual data: Language models should be fine-tuned to the specific language they are going to be used in, to help increase accuracy and better understanding of that language’s context.
- Clinical Validation: Collaborate with healthcare professionals to validate the clinical relevance and accuracy of LLMs-based diagnosis tools, to understand their perspectives on integrating such tools into real-world medical practices.
- Ethical Considerations: Address ethical considerations related to patient data privacy, especially Personally Identifiable Information (PII) that LLMs will have access to on certain tasks, and also the bias that will exist in the base model that can cause deviation to the answer.

## 6 Discussion

This review highlights the potential benefits of utilizing Large Language Models (LLMs) in the healthcare sector in Thailand. Notably, existing studies have primarily utilized LLMs for classification tasks, with limited exploration of their potential for conversational interactions with patients. This bias may be attributed to the concerns surrounding LLMs, which need to be addressed.

Once the limitations of LLMs are mitigated, they can offer numerous benefits in the healthcare sector. For instance, LLMs can help alleviate the issues of slow traffic, long wait times, and accessibility barriers that often discourage patients from seeking medical attention. By enabling patients to initially diagnose their symptoms and engage in conversational interactions with chatbots, LLMs can provide personalized advice and guidance, potentially reducing the need for human verification.

Furthermore, LLMs can be integrated into healthcare facilities to enhance workflow efficiency, such as in inpatient diagnosis coding [12]. The emergence of multimodal LLMs, capable of processing text, visual, and auditory information, offers additional opportunities for diagnosis, such as visual symptom analysis and auditory examinations.

Overall, LLMs and Large Multimodal Models (LMMs) have the potential to revolutionize healthcare by providing accurate and natural language interactions.

However, this review also highlights several limitations and challenges that need to be addressed. The reliance of LLMs on training data can introduce biases and inaccuracies, particularly in lower-resource languages like Thai. Moreover, ethical concerns surrounding the use of LLMs in sensitive fields like healthcare necessitate careful examination and weighing of benefits and concerns before their widespread adoption. It is essential to examine and balance the benefits and concerns of utilizing LLMs in healthcare before they can be integrated into everyday practices.

### 6.1 Limitations of contextual understanding and Interpretability

As LLMs is simply large artificial neural networks, it learned to make decisions by utilizing a fixed weight and bias values that are tuned in the training process (as called parameters). The decision it used to generate text is simply based on the statistical patterns of the text, they lack the reasoning behind the decisions they made, it means that LLMs cannot understand the subtlety of medical diagnoses which can had multiple different factors to keep in mind for diagnosis, which means that LLMs lacks the true understanding of the underlying medical concepts and reasons, and so, LLMs may produce inaccurate information when met with lower-resources information such as rare diseases or complex cases. And because LLMs utilize patterns for generation, they lack actual reasoning behind the decisions that it chose, which make it difficult for healthcare professionals to trust and validate its recommendations, which is a problem in critical decision-making scenarios where it require a clear understanding of the reasoning behind the suggested diagnoses or treatment options.

### 6.2 Lack of resources and Reliance on training data

How well LLMs will perform heavily relies on the training data, since any biases and inaccuracies that present in the data will be learned by the model. And depending on the dataset, the model could perform worse when utilizing the LLMs with other languages such as Thai, due to context differences and low resources about the language in the training data. It is crucial to train the base model with knowledge specific to the field you are utilizing it, and include the languages you are utilizing it in, and not in the fine-tuning phase, as fine-tuning can degrade the performance of certain knowledge even more from the base model [7]. One can utilize RAG to create a groundtruth information for the LLMs to reference in their responses [11], allowing for frequent update to the information without retraining the entire model, and to prevent hallucinations.

### 6.3 Impact on healthcare professionals

Even though LLMs can be a useful tool to healthcare professionals in reducing works that need to be done by human, one cannot completely rely on the output of such models, as that can potentially result in the lack of critical thinking and independent clinical judgement. The usage of LLMs must be as a tool to aid in the works, not replacing the role entirely.

### 6.4 Ethical Considerations

One must address the concerns of using LLMs to handle sensitive information such as patient information, as privacy and security of the data must be prioritized to protect sensitive health information. Proper consent mechanisms should be established to ensure that the patients are aware of their data being processed by the language models. And any ethical concerns regarding the potential liability and accountability of artificial intelligence systems in healthcare need to be addressed to avoid potential harm to patients and caused legal implications.

### 6.5 Real-world testing

Before LLMs can be used in real-world applications, there is a need for rigorous validation and real-world testing to assess its performance and reliability of the outcomes. Further evaluations and clinical trials are necessary to understand the true effectiveness of LLMs in improving diagnosis accuracy, reducing errors and biases and enhancing patient care and workflow.

## 7 Conclusions

This review highlights the potential benefits of utilizing Large Language Models (LLMs) in the healthcare field in Thailand, including mitigating issues related to patient waiting times and accessibility, improving diagnosis accuracy, and enhancing workflow efficiency. However, several limitations and concerns must be addressed before widespread adoption, including the reliance on training data, which can introduce biases and inaccuracies, the lack of resources with Thai languages, and ethical concerns related to patient privacy and security. Furthermore, LLMs lack true understanding of medical concepts and reasoning, which can lead to inaccurate information and difficulties in trusting and validating their recommendations. To overcome these challenges, rigorous validation and real-world testing are necessary to assess the performance and reliability of LLMs in healthcare applications. Additionally, addressing the limitations of contextual understanding, interpretability, and ethical considerations is crucial to ensure the effective and responsible integration of LLMs in healthcare. By acknowledging and addressing these challenges, LLMs can potentially become a valuable tool in improving healthcare outcomes and workflow efficiency in Thailand.

## 8 Future Considerations

As the potential benefits of LLMs in healthcare become increasingly evident, it is essential to address the limitations and challenges associated with their adoption. To fully realize the benefits of LLMs, future research should focus on the following areas:

1. Addressing biases and inaccuracies: Developing strategies to mitigate the impact of biased training data on LLMs’ performance, particularly in lower-resource languages like Thai, is crucial. This may involve developing more diverse and representative training datasets or implementing bias-reduction techniques, and collaborating with healthcare professionals and domain experts to help in tuning the datasets and implementing validation mechanisms.
2. Enhancing multimodal capabilities: Further research is needed to fully explore the potential of multimodal LLMs in healthcare, including the integration of visual, auditory, and other sensory information to improve diagnosis accuracy and patient outcomes.
3. Ethical considerations: The use of LLMs in healthcare raises important ethical concerns, including issues related to patient privacy, data security, and the potential for biased decision-making. Future research should prioritize the development of ethical guidelines and frameworks for the responsible deployment of LLMs in healthcare.
4. Human-LLM collaboration: To maximize the benefits of LLMs in healthcare, it is essential to develop effective strategies for human-LLM collaboration, ensuring that healthcare professionals and LLMs work together seamlessly to provide high-quality patient care.
5. Scalability and integration: Future research should focus on developing scalable and integrable LLM solutions that can be easily incorporated into existing healthcare systems, facilitating widespread adoption and minimizing disruption to healthcare services.

By addressing these key areas, we can unlock the full potential of LLMs in healthcare, improving patient outcomes, enhancing healthcare efficiency, and enhancing the healthcare sector as a whole.

## Supporting information

TeX Source

## Data Availability

All data produced in the present work are contained in the manuscript

