## Supplementary material for "Impact of Language Models on Healthcare in Thailand: Benefits, Challenges, and Future Opportunities": TeX Source: main.bib

@misc{vaswani2023attention,
	author = {Vaswani, Ashish and Shazeer, Noam and Parmar, Niki and Uszkoreit, Jakob and Jones, Llion and Gomez, Aidan N. and Kaiser, Lukasz and Polosukhin, Illia},
	year = {2023},
	title = {Attention {Is} {All} {You} {Need}},
}


@article{Sinha2023_hw,
	author = {Sinha, Ranwir K and Deb Roy, Asitava and Kumar, Nikhil and Mondal, Himel},
	journal = {Cureus},
	number = {2},
	year = {2023},
	month = {2},
	pages = {e35237},
	publisher = {Springer Science},
	title = {Applicability of {ChatGPT} in assisting to solve higher order problems in pathology},
	volume = {15},
}


@article{muftic_exploring_2023,
	author = {Mufti{\' c}, Fatima and Kaduni{\' c}, Merjem and Mu{\v s}inbegovi{\' c}, Almina and Almisreb, Ali Abd},
	journal = {Southeast Europe Journal of Soft Computing},
	doi = {10.21533/scjournal.v12i1.252},
	issn = {2233--1859},
	number = {1},
	year = {2023},
	month = {5},
	note = {Number: 1},
	pages = {13--41},
	title = {Exploring {Medical} {Breakthroughs}: A {Systematic} {Review} of {ChatGPT} {Applications} in {Healthcare}},
	url = {http://inquiry.ius.edu.ba/index.php/scjournal/article/view/252},
	howpublished = {http://inquiry.ius.edu.ba/index.php/scjournal/article/view/252},
	volume = {12},
}


@article{berrios1996pseudohallucinations,
	author = {Berrios, G. E. and Dening, T.},
	journal = {Psychological Medicine},
	year = {1996},
	pages = {753--763},
	title = {Pseudohallucinations: a conceptual history},
	volume = {26},
}


@misc{mckenna2023sources,
	author = {McKenna, Nick and Li, Tianyi and Cheng, Liang and Hosseini, Mohammad Javad and Johnson, Mark and Steedman, Mark},
	year = {2023},
	title = {Sources of {Hallucination} by {Large} {Language} {Models} on {Inference} {Tasks}},
}


@misc{pipatanakul2023typhoon,
	author = {Pipatanakul, Kunat and Jirabovonvisut, Phatrasek and Manakul, Potsawee and Sripaisarnmongkol, Sittipong and Patomwong, Ruangsak and Chokchainant, Pathomporn and Tharnpipitchai, Kasima},
	year = {2023},
	title = {Typhoon: Thai {Large} {Language} {Models}},
}


@misc{ghosh2024closer,
	author = {Ghosh, Sreyan and Evuru, Chandra Kiran Reddy and Kumar, Sonal and S, Ramaneswaran and Aneja, Deepali and Jin, Zeyu and Duraiswami, Ramani and Manocha, Dinesh},
	year = {2024},
	title = {A {Closer} {Look} at the {Limitations} of {Instruction} {Tuning}},
}


@misc{Gerganov_llama_cpp,
	author = {Gerganov, Georgi},
	year = {2023},
	title = {llama.cpp},
	url = {https://github.com/ggerganov/llama.cpp},
	howpublished = {https://github.com/ggerganov/llama.cpp},
}


@misc{chroma_core_chroma,
	author = {Team, Chroma Core},
	year = {2022},
	title = {Chroma},
	url = {https://github.com/chroma-core/chroma},
	howpublished = {https://github.com/chroma-core/chroma},
}


@misc{Dilocker_Weaviate,
	author = {Dilocker, Etienne and van Luijt, Bob and Voorbach, Byron and Hasan, Mohd Shukri and Rodriguez, Abdel and Kulawiak, Dirk Alexander and Antas, Marcin and Duckworth, Parker},
	title = {Weaviate},
	url = {https://github.com/weaviate/weaviate},
	howpublished = {https://github.com/weaviate/weaviate},
}


@misc{ovadia2024finetuning,
	author = {Ovadia, Oded and Brief, Menachem and Mishaeli, Moshik and Elisha, Oren},
	year = {2024},
	title = {Fine-{Tuning} or {Retrieval}? {Comparing} {Knowledge} {Injection} in {LLMs}},
}


@article{app13169450,
	author = {Suvirat, Kerdkiat and Tanasanchonnakul, Detphop and Chairat, Sawrawit and Chaichulee, Sitthichok},
	journal = {Applied Sciences},
	doi = {10.3390/app13169450},
	issn = {2076-3417},
	number = {16},
	year = {2023},
	title = {Leveraging {Language} {Models} for {Inpatient} {Diagnosis} {Coding}},
	url = {https://www.mdpi.com/2076-3417/13/16/9450},
	howpublished = {https://www.mdpi.com/2076-3417/13/16/9450},
	volume = {13},
}


@article{Munthuli_2023,
	author = {Munthuli, Adirek and Pooprasert, Pakinee and Klangpornkun, Nittayapa and Phienphanich, Phongphan and Onsuwan, Chutamanee and Jaisin, Kankamol and Pattanaseri, Keerati and Lortrakul, Juthawadee and Tantibundhit, Charturong},
	journal = {PLOS ONE},
	doi = {10.1371/journal.pone.0283095},
	editor = {V E, Sathishkumar},
	issn = {1932-6203},
	number = {3},
	year = {2023},
	month = {3},
	pages = {e0283095},
	publisher = {Public Library of Science (PLoS)},
	title = {Classification and analysis of text transcription from {Thai} depression assessment tasks among patients with depression},
	url = {http://dx.doi.org/10.1371/journal.pone.0283095},
	volume = {18},
}


@inproceedings{10202096,
	author = {Udomnuchaisup, Peachyasitt and Imsombut, Aurawan and Suwannahitatorn, Picha and Saethang, Thammakorn},
	booktitle = {2023 20th {International} {Joint} {Conference} on {Computer} {Science} and {Software} {Engineering} ({JCSSE})},
	doi = {10.1109/JCSSE58229.2023.10202096},
	issn = {2642-6579},
	year = {2023},
	month = {6},
	pages = {281--284},
	title = {Analysis of the 5Rs in {Thailand} {Medication} {Error} {Classification} through {Natural} {Language} {Processing}},
}


@article{NORASET2022104145,
	author = {Noraset, Thanapon and Chatrinan, Krittin and Tawichsri, Tanisa and Thaipisutikul, Tipajin and Tuarob, Suppawong},
	journal = {Journal of Biomedical Informatics},
	doi = {https://doi.org/10.1016/j.jbi.2022.104145},
	issn = {1532-0464},
	year = {2022},
	pages = {104145},
	title = {Language-agnostic deep learning framework for automatic monitoring of population-level mental health from social networks},
	url = {https://www.sciencedirect.com/science/article/pii/S1532046422001587},
	howpublished = {https://www.sciencedirect.com/science/article/pii/S1532046422001587},
	volume = {133},
}


@article{ponthongmak_icd10,
	author = {Ponthongmak, Wanchana and Thammasudjarit, Ratchainant and McKay, Gareth and Attia, John and Theera-Ampornpunt, Nawanan and Thakkinstian, Ammarin},
	journal = {Informatics in Medicine Unlocked},
	doi = {10.1016/j.imu.2023.101227},
	year = {2023},
	month = {3},
	pages = {101227},
	title = {Development and external validation of automated {ICD}-10 coding from discharge summaries using deep learning approaches},
	volume = {38},
}


@inproceedings{10321968,
	author = {Suvirat, Kerdkiat and Chairat, Sawrawit and Horsiritham, Kanakorn and Kongkamol, Chanon and Chaichulee, Sitthichok},
	booktitle = {2023 15th {Biomedical} {Engineering} {International} {Conference} ({BMEiCON})},
	doi = {10.1109/BMEiCON60347.2023.10321968},
	issn = {2473-7607},
	year = {2023},
	month = {10},
	pages = {1--5},
	title = {De-identification of {Thai} {Free}-text {Clinical} {Notes}},
}


@article{Boonrit2023,
	author = {Boonrit, Nantaphong and Hopkins, Ashley M. and Ruanglertboon, Warit},
	journal = {Thai Journal of Pharmacy Practice},
	issn = {3027-6136},
	number = {4},
	year = {2023},
	month = {9},
	note = {Original work published in Thai},
	pages = {1134--1148},
	title = {Evaluation of the {Appropriateness} of {Thai}-language {Answers} from {ChatGPT} for {Questions} {About} {Medication} {Use} in {Common} {Illnesses}},
	url = {https://he01.tci-thaijo.org/index.php/TJPP/article/view/264517},
	howpublished = {https://he01.tci-thaijo.org/index.php/TJPP/article/view/264517},
	volume = {15},
}
