## Supplementary material for "Impact of Language Models on Healthcare in Thailand: Benefits, Challenges, and Future Opportunities": TeX Source: my-document.pdf

After completing the eligibility assessment during the selection process (n=7), we then proceed to review the articles as follows.

### *4.3 Results*

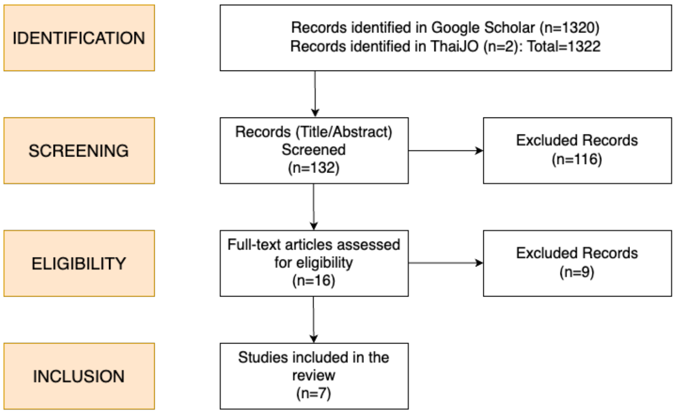


Figure 1: Flowchart of the record selection process based on PRISMA guidelines.

Table 1: Critical Analysis of usage of LLMs application in healthcare in Thailand

| Author | Study Title | Area/Focus | Outcomes | Challenges |
|---|---|---|---|---|
| Suvirat et al. [12] | Leveraging Language Models for Inpatient Diagnosis Coding | The study explores the application of language models in inpatient diagnosis coding using clinical text data that combines both English and Thai words | The study highlights the ability to capture relevant associations, and strong performance with both Thai and mixed English-Thai clinical notes. The study highlights the need for larger datasets to handle rarer diseases, while keeping the document length down as longer documents could pose a challenge to the model. | LLMs have limited context window, in which they cannot handle documents larger than certain length, and the rarity of certain diseases poses a challenge for accurate coding due to lack of information in the training data. |
| Munthuli et al. [13] | Classification and analysis of text transcription from Thai depression assessment tasks among patients with depression | The study explores the use of Encoder-Decoder transformer LLM such as XLM-RoBERTa for classification of depression by utilizing the text transcriptions of speech responses and Thai depression assessment questions | XLM-RoBERTa has a tendency to perform well in classification tasks, while only using small amount of data. Showing potential in helping with insufficient health care workers's problem and improve healthcare service accessibility. | The study concluded that speech-to-text transcription can be used for screening depression, but may not be effective for other psychiatric conditions or conditions that involve overlapping depressive symptoms. |
| Udomnuchaisup et al. [14] | Analysis of the 5Rs in Thailand Medication Error Classification through Natural Language Processing | The study explores the usage of pre-trained Large Language Models (PLMs) to enhance the detection and prevention of medication errors in hospitals, specifically expanding the coverage of the Five Rights (5R) error categorization system. | The supervised learning classification models that utilized sentence embeddings, still outperform zero-shot classification from PLMs due to insufficiency of pre-trained language models that leverage Thai language datasets from the general domain. | The study mentions the lack of PLMs employing Thai medical datasets and Thai context in general, within training. Which caused the model to underperform with Thai languages and even more so in medical-related context. |
| Noraset et al. [15] | Language-agnostic deep learning framework for automatic monitoring of population-level mental health from social networks | The researchers present LAPoMM as a framework for monitoring real-time mental health indicators from social media data without labeled datasets in low-resource languages. | LAPoMM leverages the capabilities of fine-tuned, language-independent, LaBSE model, to conduct mental signal classification and showed that extracted mental signals are in line with population-level mental health statistics | The study mentioned the lack of ability to deal with messages that are misinformed, inorganic, or automatically created, that caused the extracted social signals to be interpreted incorrectly. |

| Author | Study Title | Area/Focus | Outcomes | Challenges |
|---|---|---|---|---|
| Ponthongmak et al. [16] | Development and external validation of automated ICD-10 coding from discharge summaries using deep learning approaches | The aim of the study is to develop an automated international classification of diseases (ICD) coding tool using NLP and discharge summary texts from Thailand utilizing model trained on long English clinical text translated from Thai-English text data. | The CNN-PubMedBERT model performed second-best to PLM-ICD model, both performed well in the prediction of ICD-10 classification. | The study mentions that further external validation would be required before consideration as a clinically useful tool as Area Under the Precision-Recall Curve (AUPRCs) for these algorithms are still very modest and not sufficiently high enough for clinical use. |
| Suvirat et al. [17] | De-identification of Thai Free-text Clinical Notes | The research aims to explore, develop, and evaluate a method for de-identifying Thai free text clinical notes using language models. | The research utilized MEDPSU-RoBERTa, Multilingual BERT and WangchanBERTa, revealing that MEDPSU-RoBERTa performed best at highest score of all three models for de-identification task. | The authors present a promising results on the use of BERT architecture model for de-identification task. However, there can be other ethical requirements that needs to be fulfilled from utilizing LLMs to handle Personally Identifiable Information. |
| Boonrit et al. [18] | Evaluation of the Appropriateness of Thai-language Answers from ChatGPT for Questions About Medication Use in Common Illnesses | The study focused on validating the outputs from ChatGPT for answering questions related to medicines used for common sickness in Thai language. | The study shows that the responses were quite appropriate with some answers providing inappropriate information. | ChatGPT may respond in a way to make people think that they need to take certain medications, when in reality, they don't, this is considered a self-diagnosis and can be harmful. |

By addressing these key areas, we can unlock the full potential of LLMs in healthcare, improving patient outcomes, enhancing healthcare efficiency, and enhancing the healthcare sector as a whole.

[5] Nick McKenna, Tianyi Li, Liang Cheng, Mohammad Javad Hosseini, Mark Johnson, and Mark Steedman. Sources of Hallucination by Large Language Models on Inference Tasks, 2023.

[6] Kunat Pipatanakul, Phatrasek Jirabovonvisut, Potsawee Manakul, Sittipong Sripaisarnmongkol, Ruangsak Patomwong, Pathomporn Chokchainant, and Kasima Tharnpipitchai. Typhoon: Thai Large Language Models, 2023.

[7] Sreyan Ghosh, Chandra Kiran Reddy Evuru, Sonal Kumar, Ramaneswaran S, Deepali Aneja, Zeyu Jin, Ramani Duraiswami, and Dinesh Manocha. A Closer Look at the Limitations of Instruction Tuning, 2024.

[8] Georgi Gerganov. llama.cpp. https://github.com/ggerganov/llama.cpp, 2023. URL https://github.com/ggerganov/llama.cpp.

[9] Chroma Core Team. Chroma. https://github.com/chroma-core/chroma, 2022. URL https://github.com/chroma-core/chroma.

[10] Etienne Dilocker, Bob van Luijt, Byron Voorbach, Mohd Shukri Hasan, Abdel Rodriguez, Dirk Alexander Kulawiak, Marcin Antas, and Parker Duckworth. Weaviate. https://github.com/weaviate/weaviate. URL https://github.com/weaviate/weaviate.

[11] Oded Ovadia, Menachem Brief, Moshik Mishaeli, and Oren Elisha. Fine-Tuning or Retrieval? Comparing Knowledge Injection in LLMs, 2024.
