## Supplementary figures and images for "Impact of Language Models on Healthcare in Thailand: Benefits, Challenges, and Future Opportunities"

### prisma-a123e01b8bd35d91ecfa302b8adfd1a2.png

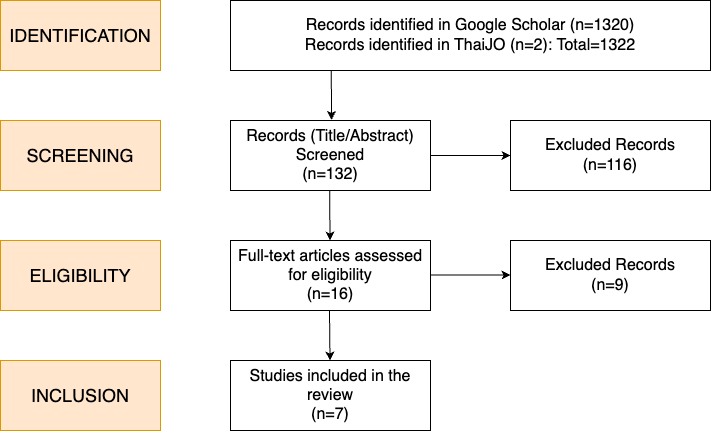
